# Transformer-based long-term predictor of subthalamic beta activity in Parkinson’s disease

**DOI:** 10.1101/2024.11.25.24317759

**Authors:** Salvatore Falciglia, Laura Caffi, Claudio Baiata, Chiara Palmisano, Ioannis Ugo Isaias, Alberto Mazzoni

**Author notes:** These authors contributed equally to this work (Ioannis Ugo Isaias, Alberto Mazzoni).

## Abstract

Deep brain stimulation (DBS) of the subthalamic nucleus (STN) is a mainstay treatment for patients with Parkinson’s disease (PD). The adaptive DBS approach (aDBS) modulates stimulation, based on the power in the beta range ([12 30] Hz) of STN local field potential, aiming to follow the patient’s clinical state. Control of aDBS relies on identifying the correct thresholds of pathological beta power. Currently, in-person reprogramming sessions, due to changes in beta power distribution over time, are needed to ensure clinical efficacy. Here we present LAURA, a Transformer-based framework predicting the nonlinear evolution of subthalamic beta power up to six days in advance, based on the analysis of chronic recordings. High prediction accuracy (> **90%**) was achieved in four PD patients with chronic DBS over months of recordings, independently from stimulation parameters. Our study paves the way for remote monitoring strategies and the implementation of new algorithm for personalized auto-tuning aDBS devices.

## 1 Introduction

Activity in the beta frequency range (12-30 Hz) in the basal ganglia circuitry is crucial for functional motor and non-motor behavior [1]. In Parkinson’s disease (PD), pathological beta power observed in the local field potentials (LFPs) of the subthalamic nucleus (STN) is associated with akinetic-rigid symptoms [2, 3], making subthalamic beta activity a promising biomarker of the patient’s motor performance [4].

Deep Brain Stimulation (DBS) is a well-established invasive neuromodulation therapy currently approved for PD, Essential tremor, Dystonia and Obsessive-compulsive disorder. Conventional DBS (cDBS) delivers a static (i.e., with constant parameters) stimulation to modulate dysregulated neural circuitry [5]. Increasing evidence suggest a possible benefit also for other brain disorders, such as epilepsy, Tourette syndrome, depression, and dementia [6].

Recently, implantable pulse generators (IPG) for DBS started to implement bidirectional neural interfaces able to both sense and stimulate deep brain areas. This evolution is opening new possibilities for understanding the pathophysiology of brain disorders and discovering methods for disrupting, normalizing or reversing it by means of electrical stimulation [7]. From this, adaptive DBS (aDBS) was conceived as a new type of DBS, introducing a closed-loop delivery of stimulation aimed to follow the clinical state of the patient based on the beta power extracted from local field potential (LFP) recordings in STN [8, 9]. The currently available aDBS algorithms for PD are based on patient-specific beta power thresholds. The Percept PC/RC device (Medtronic Inc) can operate in a single-threshold or dual-threshold mode. The single threshold approach suppresses beta activity when it exceeds a certain level for a given time10 with a rapid current ramping. The double-threshold algorithm instead confines beta oscillations between two beta power thresholds [10] delivering a slow modulation of the delivered current. In contrast, the AlphaDBS device (Newronika SpA), used in this study, provides a linear modulation of the stimulation current within two beta power thresholds [11–13] (Figure 1a). With this algorithm, stimulation is constantly modulated between the two beta power thresholds [12].

**Fig. 1:**
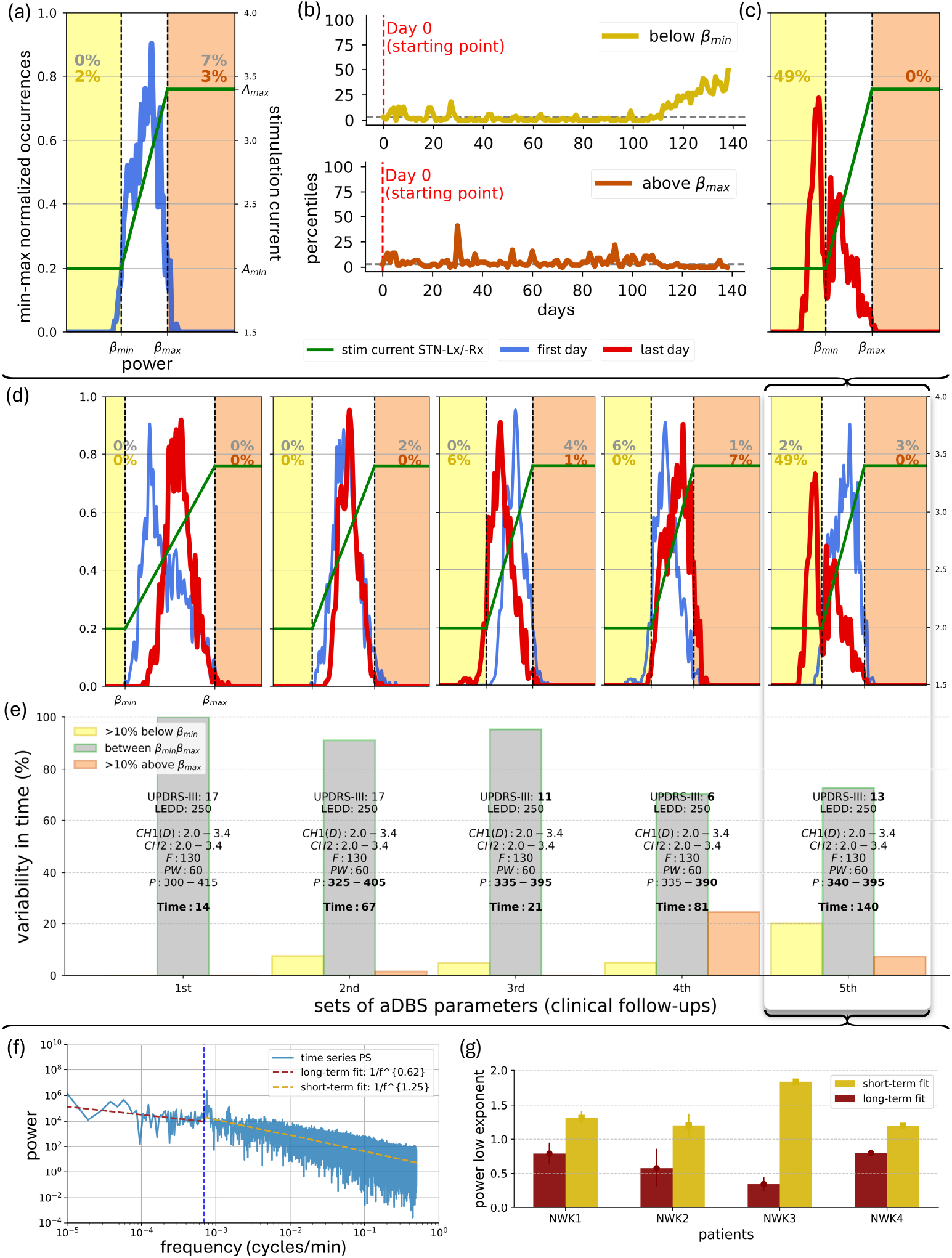
Explorative Data Analysis (EDA). (a) The aDBS parameters are set (day-0) based on the clinical condition of one patient (NWK1) and the daily distributions of subthalamic beta-power from previous days (blue). Profile of the delivered current (green-line; in Amin, Amax or modulated with the linear proportional algorithm). *β*_*min*_ and *β*_*max*_ (dashed-vertical-lines). Daily percentages below and above these thresholds (yellow and orange, respectively) and prior-to-recalibration (grey). Distributions presented as min-max normalised occurrences over beta-power, resampled, and averaged over each successive pair-of-bins. (b) Variability profiles for NWK1 over time assessed for the distribution percentiles below-above the two beta thresholds. (c) Distribution of patient-specific (NWK1) subthalamic beta-power of the last day (day-140) before a subsequent visit for aDBS reprogramming. (d) Same as (a, c) but combined for every visit (each box is a visit). (e) Variability was assessed within each patient’s set of aDBS parameters used, distinguishing between net shifts of the beta distributions (panel d), expressed in fractions of time, below *β*_*min*_ (dark-yellow) leading to understimulation, above *β*_*max*_ (dark-orange) leading to overstimulation, and between the two thresholds (red). UPDRS-III: Unified Parkinson’s Disease Rating Scale motor part score (maximum score is 108) - value assessed on the morning of the clinical-follow-up. LEDD: drug dosage (mg) as Levodopa Equivalent Daily Doses [33] - drug dosage valid for the period prior-to the corresponding follow-up. CH1: left hemisphere; CH2: right hemisphere; D: driver channel/hemisphere. List of new set-of-aDBS-parameters valid until the next follow-up; Numbers after CH1 and CH2 refer to Amin: stimulation amplitude (mA) delivered at or below *β*_*min*_ and Amax: stimulation amplitude (mA) delivered at or above *β*_*max*_; F: frequency-of-stimulation (Hz); PW: pulse-width (*µs*); P: *β*_*min*_-*β*_*max*_, lower-upper beta-power thresholds for current adjustments. Time: number-of-days for which the new set-of-aDBS-parameters apply. (f) Power-spectrum (PS) of the recorded time series. Short- and long-term evolutions within the PS are fitted separately to a linear power-low model (yellow and red, respectively). The cut-off point was fixed at 24h (1440min) to account for circadian rhythms. (g) Variability of the power-low exponent in the high-frequency and low-frequency fit of the patients’ PS according to their different set-of-aDBS-parameters for all four patients individually.

Overall, aDBS can be viewed as a closed-loop control system, in which the process output (LFPs) is continuously monitored and fed back into the controller (IPG) to adjust the input (stimulation current amplitude) [4, 10]. In the present configuration of the aDBS workflow, the STN is the process, the LFPs are the output, and the aDBS device is the controller. The most difficult component of programming in aDBS is setting beta thresholds to maintain the adaptive component of aDBS over time and avoid periods of over-stimulation or under-stimulation, which might lead to long-term adverse events [14–16] or suboptimal symptom improvement. Indeed, patients undergoing aDBS therapy require periodic retuning of algorithm parameters. The difficulty is thought to be mainly related to beta modulation by dopaminergic drugs, and its physiological role in the execution of multiple motor and non-motor tasks [17–19]. An additional critical aspect is the impact of stimulations on the long-term evolution of beta oscillations. Algorithms are needed that can detect and predict the evolution of subthalamic beta oscillations for automatic recalibration of DBS parameters, especially beta thresholds. The adoption of these algorithms holds the key for novel restorative therapies.

A number of deep learning (DL) architectures have been developed with the specific purpose of addressing time-series forecasting [20]. The translation of such solutions to the field of PD and brain disorders presents several challenges. These include the large amount of chronic recordings [21] from a single patient, collected over months or years [12], as well as algorithms for the identification of the hidden patterns of evolution of the driver neural signal [22]. Model-based approaches dealing with the prediction of dynamic responses of neuromodulation are limited in their ability to consider the full range of stimulation parameters [23]. Machine learning algorithms have been trained for seizure detection monitoring scalp electroencephalography and electrocardiography from the same epileptic patients over years [24, 25]. Adaptive Bayesian optimization has been studied to suggest optimal recalibration in amputees implanted with neural interfaces [26]. Similarly, Bayesian optimization of vagus nerve stimulation has been used for optimizing neural stimulation parameters [27]. Furthermore, several studies addressed short-term forecasting of deep brain activity [28, 29]. However, no studies have been yet conducted for neurophysiological long-term forecasting [30–32] of the activity of deep brain areas to optimize neuromodulation strategies.

In this study, we propose LAURA (Learning betA-power distribUtions through Recurrent Analysis), a Transformer-based framework designed to forecast the long-term evolution of subthalamic beta power activity under aDBS therapy in PD. The framework will allow to train a patient-specific algorithm for each individual patient able to predict in a personalized way beta power drifts and fluctuations over long timescales based on a limited temporal interval of recordings.

## 2 Results

### 2.1 Study of long-term STN beta power evolution over months on chronic recordings for personalized DBS device tuning

We investigated the long-term evolution of subthalamic beta power in patients with PD and chronic STN DBS (see 4.Methods). We aimed to define an algorithm capable to predict the evolution of beta activity as a key aspect of novel autotuning IPGs. All patients (n=4) showed significant and sustained clinical improvement with bilateral STN-DBS (range 76-90% reduction in UPDRS-III score comparing active stimulation with immediate off stimulation). All patients underwent both aDBS and cDBS (4.Methods). All preferred aDBS except one (NWK4), who chose cDBS stimulation at the last follow-ups for better tremor suppression. In this patient, we noted the reappearance and habituation of tremor to amplitudes of stimulation toward Amin and the need to maintain a small adaptive therapeutic window (Amin-Amax gap). The UPDRS-III scores and the LEDD [33] results are provided as supplementary data (Supplementary Figure 1).

### 2.2 STN beta power distributions show large linearly uncorrelated fluctuations over several days

We observed that the beta power distribution referred to set the stimulation thresholds (Figure 1a and 4.Methods) drifts over time (Figure 1b). This eventually leads to a shift in the distribution, finally displaying a relevant fraction of time spent with a beta power below *β*_*min*_ (as in Figure 1c) or above *β*_*max*_. This in turns leads in the first case to an understimulation, i.e., to long periods of time spent with minimal and subeffective stimulation, and in the second case to an overstimulation, i.e., long periods of time spent with maximal stimulation. This occurs for every recalibration of the aDBS parameters (i.e., every aDBS setting; Figure 1d, 1e and Supplementary Figure 1). We evaluated then the characteristic timescale of the drifts. The spectral analysis (Figure 1f) demonstrated that on short time scales (i.e., shorter than 24h) the evolution of beta power exhibited an exponent greater than 1 for all patients (Figure 1g). In contrast, over timescales longer than a day, the evolution of beta power exhibited a lower exponent (Figure 1g), indicating the absence of linear correlations, and large fluctuation in the time domain. Notably, such behaviour does not depend on variations over time of the beta peak in the spectrum (Supplementary Figure 2). This shows that beyond the within day fluctuations, that are efficiently managed by the standard aDBS algorithm, there are large uncorrelated fluctuation on the temporal scale of several days.

**Fig. 2:**
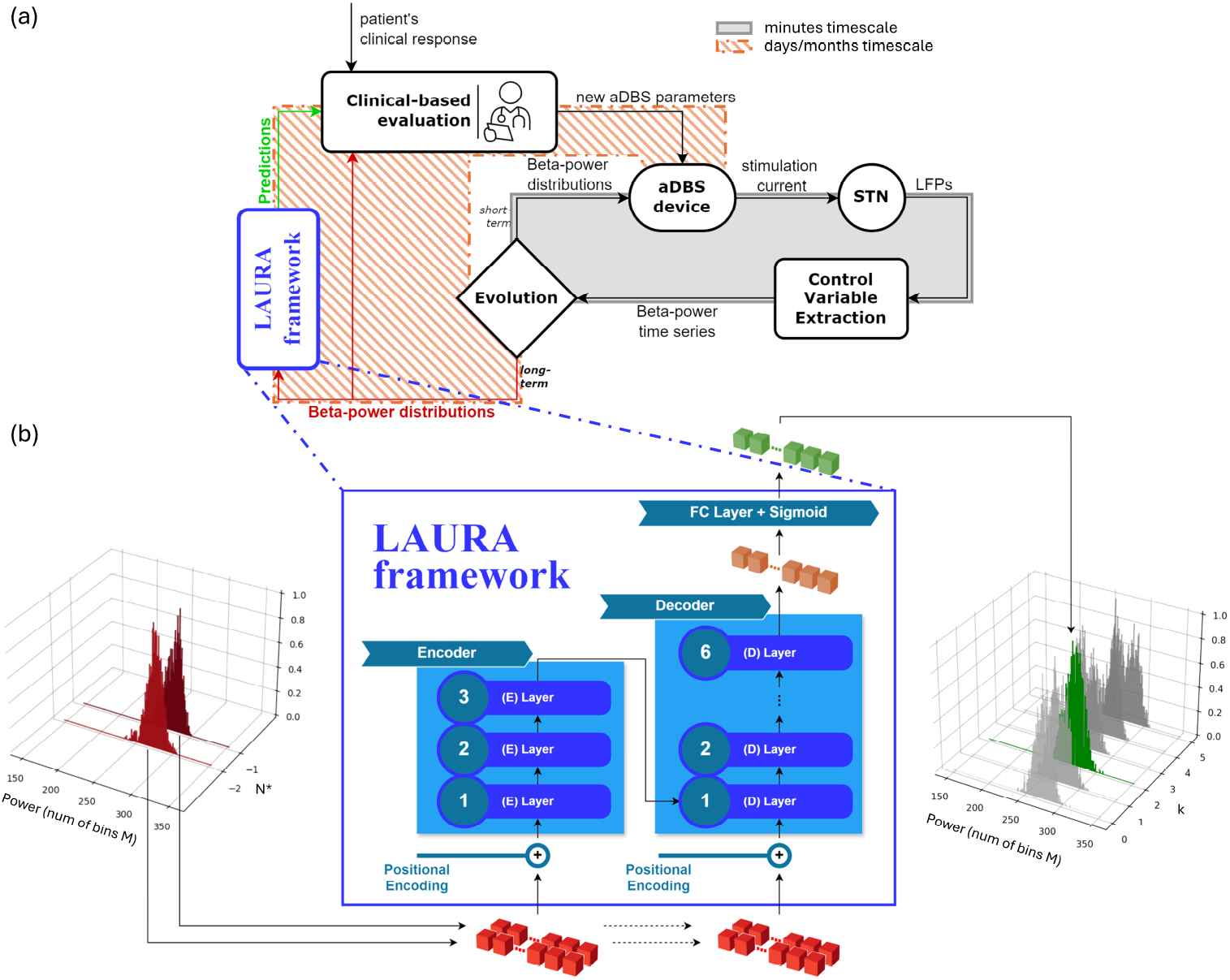
LAURA framework within the aDBS parameters tuning strategy. Learning betA-power distribUtions through Recurrent Analysis (LAURA) as a framework within the aDBS workflow to improve long-term DBS programming strategy for PD. (a) Block diagram representing aDBS as a closed-loop control system in which the process output (local field potentials, LFPs) is continuously monitored and fed back into the controller (IPG) to adjust the input (stimulation current amplitude). In the present configuration, the STN is the process, the LFPs are the output, and the aDBS device is the controller. The control loop is composed of two algorithms acting on two separated timescales. On the timescale of minutes (i.e., short-term evolution), the modulation is changed according to the dynamical symptom-related fluctuations of the beta power through the standard aDBS proportional algorithm linking beta power and stimulus current (solid box). On the timescale of days/months (i.e., long-term evolution), the parameters of the fast aDBS algorithm are updated based on the expected drifts of the whole daily beta distribution through LAURA algorithm combined by the neurologist clinical evaluations (dashed box). (b) LAURA framework consisting of a Transformer model with 3 encoder layers, 6 decoder layers, and a FC layer with a sigmoid function as output non-linearity. It takes in input a sequence of *N* ^*^ daily-distributions (red), where *N* ^*^ is the optimal number of past days required to perform a prediction, captures the model’s understanding of the entire sequence (orange), and provides as output one daily distribution (green) among the first one up to the sixth one succeeding the input distributions (*k* ∈ [0, 5]). Distributions are expressed with number of bins M. Here *N* ^*^ = 2, *M* = 206, *k* = 2.

### 2.3 LAURA: a personalized Transformer-based framework for forecasting STN beta power distribution

In this scenario, we propose a deep learning framework (LAURA) designed to predict, for each individual patient with a personalized algorithm, the slow shifts in the sub-thalamic beta power distribution across days, based on previous recording history of the patient. This long term framework will be integrated with the current fast aDBS algorithm taking care of the symptom related fluctuations (Figure 2a). LAURA’s prediction of beta distirbution drifts, combined with clinical evaluation of the patient condition, will support an effective recalibration of aDBS parameters on a daily basis (Figure 2a). Briefly, LAURA algorithm is a Transformer model with three encoding layers and six decoding layers (Figure 2b, 4.Methods). It receives as input the past distributions as a matrix N x M where N is the number of days in the past the prediction is based on, and M is the binning of the beta power distribution of each day. It delivers as output a vector with dimension M with the predicted distribution of K=k+1 days in the future (where k is the index of the daily distributions list; Figure 2b).

We evaluated, for each patient individually, the performance of LAURA(N,K) algorithms trained and validated on datasets varying N and K (Supplementary Figure 3) for several seeds, while keeping M=206 for simplicity (see 4.Methods). As a criterion, we computed the weighted mean absolute percentage error (wMAPE) between the true and predicted distributions to account for the different variations in the values along dimension M within the distribution (4.Methods).

**Fig. 3:**
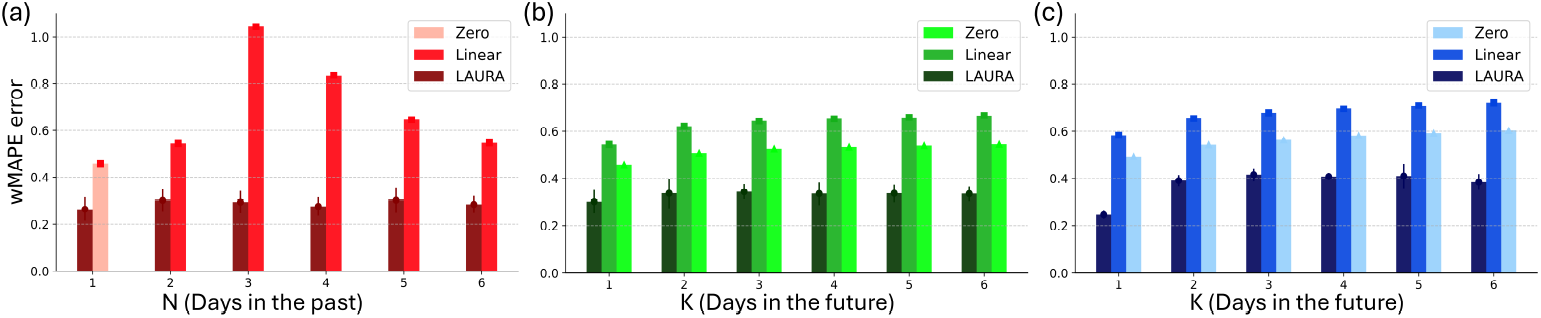
LAURA’s patient-dependent performance. LAURA performance on patient NWK1 assessed by monitoring the weighted mean average percentage error (wMAPE) between the observed and predicted distributions (see Suppl. Figure 4 for all patients) as a similarity metric (4.Methods). (a) One-step-ahead prediction over a single aDBS setting. The prediction error is monitored for six different length (from 1 to 6 daily distributions) of the input sequence. The wMAPE of LAURA (dark red) visualized against the wMAPE of a deterministic linear regressor (red), collapsing to a zero-order regressor (light red) when the input sequence has length 1 sample. LAURA outperforms both the zero-order regressor when based on only 1-day of history, and the linear regressor when increasing the length of the history sequence. The zero and first-order regressors show a deterioration in forecasting performance when the length of the history sequence is increased from 1 to 3 daily distributions, and then an improvement in performance towards convergence with the results obtained with a history of only 1-2 days by increasing the length of the sequence. The forecasts of LAURA show quantitative and qualitative improvements independent of the length of the history sequence on which the forecast is based. (b) Multi-step-ahead prediction over a single aDBS setting. The wMAPE is monitored on predictions from 1 up to 6 days ahead, based on the patient’s history sequence of optimal length *N* ^*^(=2 days, for patient NWK1) found experimentally. The wMAPE of LAURA (dark green) visualized against the wMAPE of a linear regressor (green) and the wMAPE of a zero-order regressor (light green). LAURA outperforms both zero and first-order regressors in multi-step-ahead prediction. While linear regressors increase the error in their predictions as the prediction step-ahead increases, LAURA manages to keep the error significantly lower than the one computed by linear regressors and almost constant. (c) Multi-step-ahead prediction over multiple aDBS settings. LAURA can cope with intra-patient variability due to the recalibration of the aDBS device. The error of the predicted distributions is comparable to that obtained on one set of parameters.

We first analysed the single-setting scenario. Testing N from 1 to 6 days for K=1 (Figure 3a and Supplementary Figure 4), LAURA achieved a stable performance (wMAPE-LAURA range across N and patients [0.25 ±0.03 0.47 ±0.03]). Crucially, it outperformed linear regressors for every patient and every combination of parameters (wMAPE-Linear range across N and patients [0.40 1.36]). We then set *N* = *N* ^*^ as the shortest patient-dependent efficient sequence of past distributions (4.Methods) and tested the algorithm performance in the multi-step-ahead task varying K (Figure 3b and Supplementary Figure 4). We found that LAURA performance was stable and again outperformed linear regressors for each K (across K and patients: wMAPE-LAURA range [0.25 ±0.03 0.55± 0.07], wMAPE-Linear range [0.48 ±0.93], wMAPE-Zero range [0.40 ±0.79]). We finally tested the ability of LAURA to cope with intra-patient changes in aDBS parameters due to re-programming of the IPG (i.e., multiple-setting scenario; Figure 3c and Supplementary Figure 4). We found that LAURA displayed only a minor loss (wMAPE-LAURA range across K and patients [0.25 ±0.020.75 ±0.11]), and still outperformed linear regressors for each K (across K and patients: wMAPE-Linear range [0.52 0.95], wMAPE-Zero range [0.44 0.80]). The same analysis performed only on recordings during awake hours (9am – 9pm), did not lead to any increase in performance with respect to whole-day distributions (Supplementary Figure 5), and LAURA did still outperform linear regressors for every combination of parameters.

**Fig. 4:**
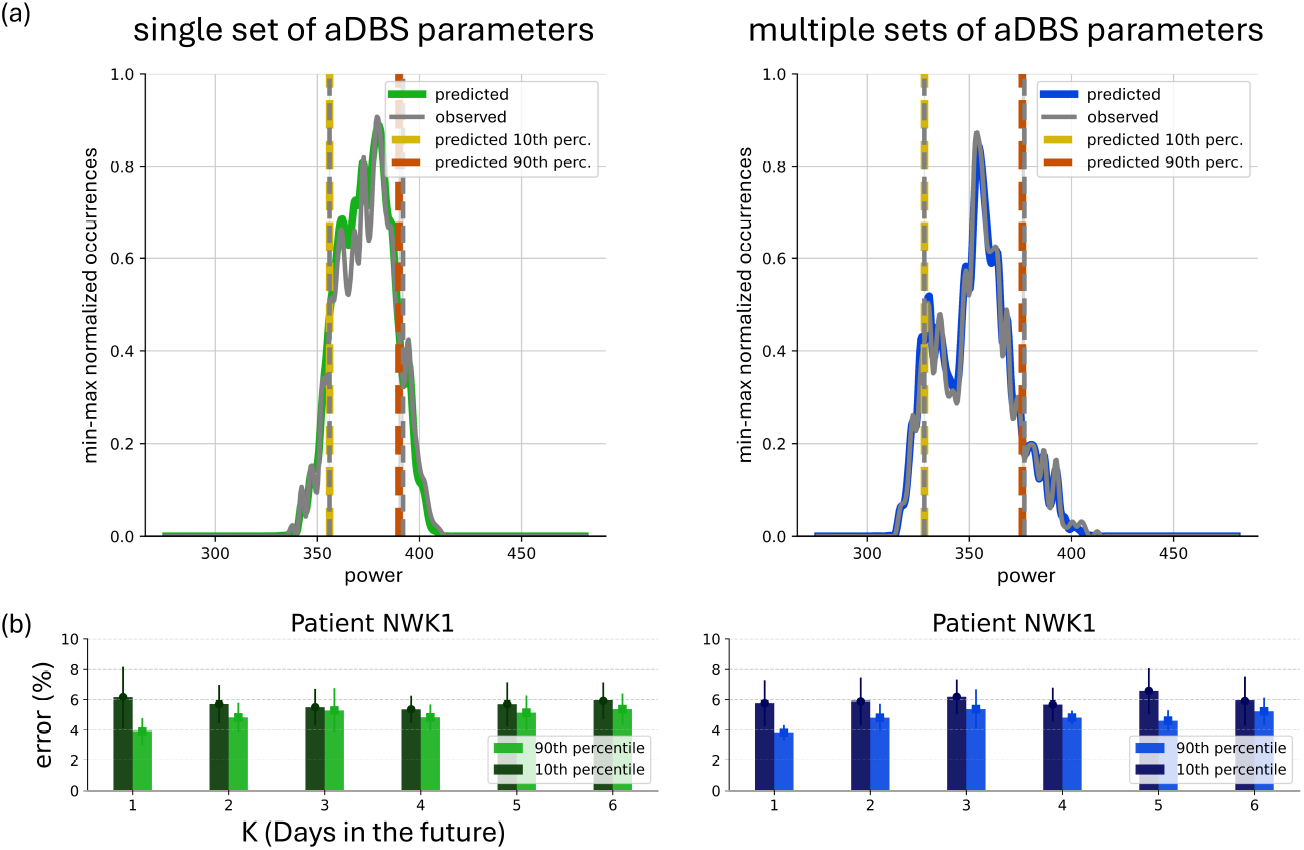
Functional evaluation of predicted distributions under aDBS setting. Evaluation of the predictions assessed by monitoring the inaccuracies of LAURA in identifying the 10th and 90th percentiles of each daily distributions on patient NWK1 (see Suppl. Figure 6 for all patients). (a) Examples of observed (grey) and predicted (green on the left, blue on the right) distributions in the task 2days →1day-ahead within both cases of single aDBS setting (left) and multiple aDBS settings (right). For the sake of clarity, the distributions have been presented as min-max normalised occurrences over beta power, by resampling and taking the average of each successive pair of bins. The 10th and 90th percentiles are reported for the observed (in grey) and the predicted (in dark yellow and dark orange, respectively) distributions. (b) Inaccuracies of LAURA in identifying the 10th (dark) and 90th (light) percentiles for patient NWK1, for both cases of single (left) and multiple (right) aDBS settings. Results support LAURA as a patient-specific system.

**Fig. 5:**
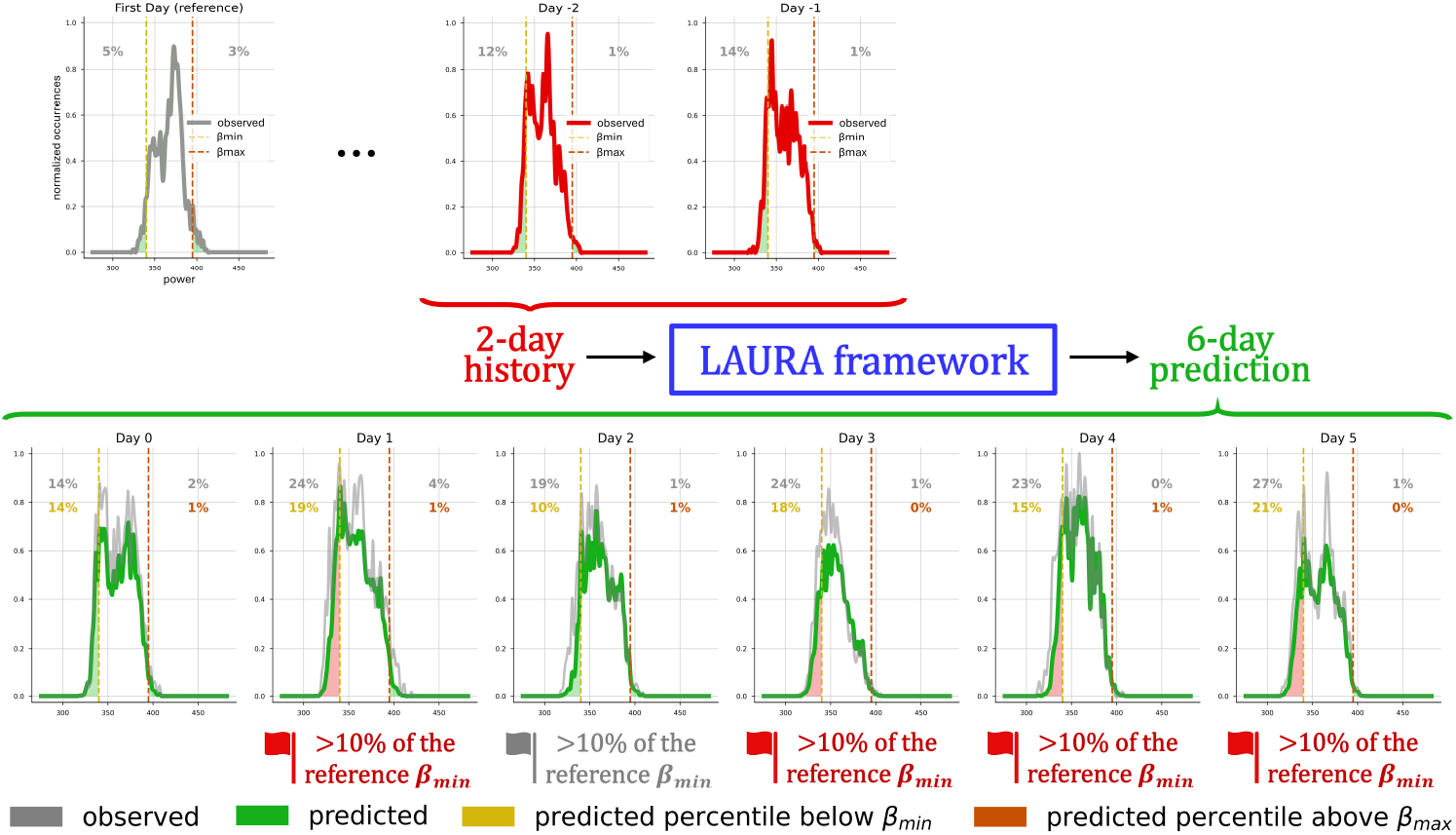
Illustration of a possible application of LAURA beta-forecast. The incorporation of LAURA into the existing aDBS programming workflow would represent a substantial advancement in the field of personalised medicine, with the potential to facilitate the widespread adoption of aDBS therapy as a long-term treatment strategy for PD. For the sake of clarity, the distributions have been presented as minimum-maximum normalised occurrences over beta power, by resampling and taking the average of each successive pair of bins. At the first visit (top-left, grey panel), the neurologist defines the optimal parameters for aDBS based on the clinical evaluation and previous subthalamic recordings. This entails defining the percentiles of the daily distribution below *β*_*min*_ and above *β*_*max*_. These two thresholds allow to define the amount of time the patient is at Amin, Amax actually linearly adapted. LAURA receives as input a sequence of *N* ^*^ (=2 for patient NWK1; day -2 and day -1, top row in red) observed daily distributions and provides as output the predicted subthalamic beta power distributions over 6 days in advance (bottom row in green), together with a reliable estimate of their tails (i.e., the percentiles of the distribution below *β*_*min*_ and above *β*_*max*_). This allows alarms to be set whenever the tails exceed a safe range (e.g., 10%) around thresholds for aDBS reprogramming and paves the way for remote monitoring strategies and the implementation of new algorithm for personalized autotuning devices.

In addition, we tested a Random Forest algorithm (RFA) to predict the evolution of the beta band and compare it with piecewise and linear order regressors. The RFA does not make any a priori assumption on the dynamics in the data, thus it is able to capture non-linearities and, at the same time, it is sufficiently different from the LAURA’s Transformer approach to make the comparison relevant (see 4.Methods). Our results reveal that adopting the RFA results in a substantial reduction in prediction error when compared to both first-order and zero-order regressors, though RFA’s error remains higher than LAURA’s (Supplementary Figure 6).

### 2.4 Functional evaluation of LAURA for the programming of aDBS devices

Finally, we computed the error in identifying the 10th and 90th percentiles in each predicted distribution as an estimation of the validity of our predictions according to the two beta thresholds (i.e., *β*_*min*_ and *β*_*max*_) for both single and multiple settings (Figure 4 and Supplementary Figure 7; 4.Methods). Very accurate results were obtained for predictions within a single set and multiple sets of aDBS parameters, with an average error in identification of both percentiles below 10% (range across K and patients of the accuracy error: single-setting scenario 10th-percentile [3.53 ± 0.34 7.88 ± 1.24] 90th-percentile [3.47 ± 0.63 9.00 ± 2.05], multiple-setting scenario 10th-percentile [2.18 ± 0.64 6.90 ± 0.64] - 90th-percentile [2.96 ± 0.57 6.41 ± 1.11]).

## 3. Discussion

We presented LAURA, a personalized Transformer-based framework for forecasting subthalamic beta power distribution and providing the clinician with a more complete picture of the patient’s STN-LFP beta power and its predicted evolution, thus facilitating informed decision-making (Figure 2 and Figure 5). Our algorithm analyses home recordings with 1-min resolution in chronically stimulated patients. LAURA proved efficacy in both the one-day-ahead and multi-day-ahead prediction tasks, with forecasts extending up to six days. The performance vastly outperformed linear algorithms. The approach was validated in four parkinsonian patients exhibiting heterogeneous subthalamic beta power dynamics and independently of stimulation parameters.

LAURA resulted from the translation of the recent achievements in natural language processing (NLP) and time-series forecasting into the domain of brain disorders through the use of deep learning (DL) techniques. The Transformer model [34] represents one of the most sophisticated DL units for sequence-to-sequence tasks, due to its internal multi-head attention (MHA) layer based on a general concept of the sparsity of interactions within the sequence. Transformers displayed superior scaling law, outperforming other generative stochastic models [35]. This enables the development of large frameworks capable of handling a diverse range of tasks, as exemplified by the latest large language models (LLMs) dealing with heterogeneous data [36]. We translated the Transformer’s approach into a DBS scenario, by considering a sequence of patientspecific subthalamic beta power distributions. Each distribution represents a token, for which we achieve a contextual embedding and perform one- and multi-step-ahead predictions (Figure 2b).

Crucially, the need of DL architectures for large sets of diverse data for training [37– 39] poses a challenge in the clinical setting. Indeed, our results rely on the availability of heterogeneous recordings collected with a resolution of minutes over one year [12]. For each patient, several consecutive days within different sets of DBS parameters have been analysed (Figure 1 and Supplementary Figure 1).

We investigated the personalised effect of chronic DBS on STN beta power and predicted its evolution in the long-term.

We first showed that this evolution is not described by a random process (Figure 1f-1g). This allowed us to further highlight the presence of non-linear long-term biological correlations. Noteworthy is that LAURA manages to generate predictions based exclusively on neural data, thereby revealing the presence of latent patterns intrinsic to subthalamic activity in response to chronic DBS. Furthermore, LAURA outperformed linear regressors in predicting the changes in beta-power distributions over a timescale of days (2.Results, Figure 3). When considering the one-day-ahead task with respect to the length of the history sequence (N), these linear regressors showed an initial deterioration in forecasting performance when N was increased up to three daily distributions (Figure 3a). As the length of the history sequence was increased, this effect was followed by an improvement in performance, which converged with the results obtained with a history of only one or two days. When considering the multi-day-ahead task, linear regressors increased the error in their predictions as the prediction-step-ahead increased (Figure 3b). Interestingly, the zero order regressor outperformed the first-order regressor, ruling out any linear evolution in the subthalamic beta activity. Further confirmation of the inherent nonlinearity of the dynamics is provided by the results of the RFA in predicting the evolution of the subthalamic beta activity, since it outperforms the zero-order regressor (Supplementary Figure 6). These findings justify our analysis of temporal sequential dependencies conducted via LAURA.

The forecasts of LAURA outperformed all other approaches and was independent from the length of the history sequence, the step-ahead in the prediction, and the DBS parameters (Figure 3c). We have also shown that LAURA can provide accurate predictions of the daily distributions, not only of their central region, but also of the tails of the distributions estimated from their 10th and 90th percentiles (Figure 4). This aspect is particularly important because it ensures that LAURA can predict the tails of the beta distribution with respect to predefined thresholds (in percentiles). With the AlphaDBS device, these thresholds allow defining the percentage of time when stimulation is delivered at the Amin, Amax or actually linearly modulated in between (4.Methods).

LAURA framework represents a significant advancement within the aDBS work-flow, potentially supporting the widespread adoption of the aDBS therapy as a long-term treatment strategy for PD. The first immediate use of LAURA is for early DBS reprogramming intervention through the configuration of automated warnings, triggered when beta power oscillations are expected to deviate more than 10% from the *β*_*min*_ and *β*_*max*_ thresholds defined during an in-person visit (Figure 5). This can be readily implemented with the AlphaDBS device by the fact that neural recordings used for the aDBS mode are continuously saved, downloaded during IPG charging, and sent to the treating neurologist via a WebBioBank [40].

Furthermore, the ability to predict the evolution of patient-specific subthalamic beta oscillatory activity is a key aspect of future personalized [41] auto-tuning IPGs. A first direct application would be the automatic readjustment of *β*_*min*_ and *β*_*max*_ to maintain stimulation in aDBS mode over time. However, the predictive aspect of LAURA is even more important for the study and development of new disease-modifying DBS paradigms to restore physiological subthalamic activity over the long term.

This study presents several limits. First of all, LAURA generates a warning about the mismatch between future beta distribution and current stimulation settings, but it does not predict an entire set of novel optimal stimulation settings. The programming adjustments proposed by LAURA pertains solely to the two thresholds *β*_*min*_ and *β*_*max*_; no indication on the minimum (Amin) and maximum (Amax) stimulation currents are provided, which for now shall remain set according to the individual clinical response (4.Methods; Figure 1, Supplementary Figure 1).

Second, the great number of DBS parameters does not allow LAURA to forecast instantaneously the evolution of subthalamic beta activity according to a new stimulation parameters, without a prior monitoring period. However, the ability of the AlphaDBS device to transfer the recorded data to a cloud system (i.e., WebBioBank [40]) ensure consistent LAURA updates and predictions. Note that while the number of DBS parameters available constitutes a limitation, the relatively small number of patients in this study does not, since we deal with intra-patient variability.

Third, clinical evaluations are only provided at follow-up appointments rather than on a daily basis. For this reason, our comprehension of the neural data is still insufficient to fully understand the observed or predicted behaviour from a clinical perspective. It is interesting to note that not all fluctuations resulted in a deterioration of the patient’s condition. To this end, we already started monitoring patients with wearable devices to better characterize the clinical correlations of subthalamic beta activity with daily fluctuations in relation to the patient’s motor and non-motor activities.

The human/clinician-in-the-loop approach [42, 43] represents today the optimal strategy for LAURA framework and it opens the way for novel long-term neuromodulation strategies aimed at restoring brain function not only for PD, but also for a variety of other neurological disorders, such as stroke, epilepsy, depression, and dementia.

LAURA also represents a step towards a deeper understanding of brain activity in patients chronically implanted for DBS. It leads to the development of a comprehensive, explainable DL architecture capable of combining neural data with chronic informative data pertaining to the clinical state that will ultimately lead to autotuning devices. The future integration of clinical diaries and wearables will result in a more complete framework that resembles the mechanisms of current LLMs [44]. This will facilitate the correlation of latent patterns of subthalamic activity with clinical outcomes and motor and non-motor activities of daily living.

## 4 Methods

### 4.1 Patients and data acquisition

This study involving humans was conducted in accordance with the Declaration of Helsinki and approved by Milano Area 2 Ethics Committee (approval: 165-2020 and 93-2023bis). The study was conducted in accordance with the local legislation and institutional requirements. The participant provided written informed consent to participate in this study. Written informed consent was obtained from the individual(s) for the publication of any potentially identifiable images or data included in this article. We recorded STN LFPs from the chronically implanted DBS electrodes (3389 leads, Medtronic Inc) in four patients with idiopathic PD (Hoehn and Yahr stage 2 of 5 in best medical treatment). All patients received STN DBS treatment for clinical needs and the AlphaDBS IPG in the context of a clinical trial [45]. The study was approved by the local Ethics Committee (approval: 165-2020 and 93-2023bis) and conformed to the declaration of Helsinki. The patients gave written consent prior to participation in the study. Demographic and clinical characteristics of the patients are listed in Supplementary Table 1.

Patients were clinically assessed with the Unified Parkinson’s Disease Rating Scale motor part (UPDRS-III) performed in the morning after the intake of the usual drug therapy. Medications are noted as Levodopa Equivalent Daily Dose (LEDD) [33].

The programming paradigm of aDBS with the AlphaDBS device has been previously described [11, 12]. In brief, the AlphaDBS device linearly modulates the stimulation current delivered to both STN based on the average STN LFP power recorded in one hemisphere, calculated in a patient-specific beta frequency range, and normalized for the total power in the 5-34 Hz range. The averaging procedure consists of an exponential moving average with a time constant of 50 s over the beta power samples calculated with 1-s resolution. The stimulation current is adjusted between two limits [Amin, Amax], clinically defined. Amin is the amperage with 40%-50% clinical benefit in meds-off state (overnight) or wearing-off state (end of levodopa benefit), and Amax is the maximum clinically effective amperage without side effects in meds-on condition. The stimulation frequency and pulse width are clinically defined and remain fixed.

The recording contact pair is chosen as the one showing a more prominent and stable frequency (beta-peak) in the beta band. Two frequencies (Lbound and Hbound) are set around the beta-peak and define a patient-specific beta range; the beta power in this range represents the input signal used to modulate the delivered current. The AlphaDBS device saves the power of the beta range every minute (1-min resolution). The distribution over time of these measurements is visualized as a histogram and is used to set the two thresholds [*β*_*min*_, *β*_*max*_]. For beta power values less than *β*_*min*_ the stimulation current delivered is Amin and for values greater than *β*_*max*_ the current delivered is Amax, between the two thresholds the stimulation current is delivered in aDBS mode (Figure 1a-1c). In terms of programming options, these thresholds allow to define the amount of time the patient is at Amin, Amax or actually linearly modulated in between (Figure 1d). In general, we set *β*_*min*_ and *β*_*max*_ to allow current modulation for *>* 70% of total awake time. Figure 1e and Supplementary Figure 1 illustrate the sets of stimulation parameters for the patients recruited in the study.

### 4.2 Explorative Data Analysis (EDA)

Beta power thresholds are the innovative and crucial aspect of programming in aDBS. Currently, with both the AlphaDBS device and Percept PC/RC, they can only be changed manually. This is an important limitation because the beta power distribution in the patient-specific range can change significantly over time, skewing toward or exceeding either threshold. This may depend on multiple factors, which are still being studied, involving PD-related, physiological and compensatory beta oscillatory activity, e.g., the evolution of PD, including as a response to chronic aDBS, changes in drug dosing, or even due to a change in lifestyle habits and activities after surgery. Currently, the timing of these changes in beta activity is not known or predictable, and the patient must be closely monitored in person, or remotely, thanks for example to WebBioBank (Newronika SpA) [40]. Of note, with significant changes in beta power distribution (an example in Figure 1), the patient is at risk of being stimulated in cDBS with a fixed current delivered in Amin or Amax, in the former case with poor clinical benefit and in the latter with the possible occurrence of adverse events from chronic overstimulation.

In this study, to provide a quantitative estimate of beta power variability, we defined the percentages of non-consecutive minutes with beta power measurements below *β*_*min*_ (in Amin), above *β*_*max*_ (in Amax), and between the two thresholds (in aDBS). We then arbitrarily defined a 10% deviation at follow-up as the cutoff for a clinically meaningful shift (Figure 1e). The value 10% means that the patient was stimulated for 2.4 h more in either Amin or Amax than set at initial programming. The variability (*σ*_*below*−*βmin*_, *σ*_*above*−*βmax*_) within a given set of aDBS parameters is therefore defined as the ratio between the number of days exceeding the tolerance time interval (*M*_*below*−*βmin*_, *M*_*above*−*βmax*_) and the number of days on which the same setting was used (*M*_*setting*_):

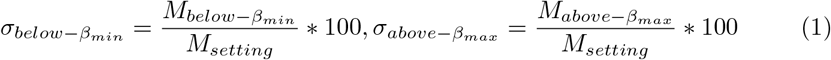

In the event of cDBS (Supplementary Figure 1d), we performed a similar analysis using the 10th and 90th percentiles of the beta power distributions.

The beta power spectrum range was extracted for each patient (Figure 1f). This spectrum is characterised by a frequency resolution of Δ*f* = 1*/T* = 1*/N*_*min*_ = 10^−5^ cycles per minute and a frequency range from 10^−5^ to the Nyquist frequency *f*_*Nyquist*_ = 0.5 cycles per minute in steps of Δ*f* cycles per minute. We divided the spectrum into two dynamics: the short-term evolution (i.e., within 24h), at higher frequencies, and the long-term evolution (i.e., over 24h), at lower frequencies. Both have been fitted to a power low model *Cf* ^−*α*^, where the fitted exponent *α* provides insight into the nature of the time series. Indeed, the three possible cases are *α*≈ 0 (white noise), indicating an (almost) uncorrelated process, where each value in the series is independent of the others; *α*≈ 1 (pink noise), indicating a balance between randomness and predictability; *α* ≈ 2 (Brownian noise or random walk noise), suggesting a system with strong temporal correlations. A flat power spectrum at low frequencies indicates the absence of long-term correlations, as opposed to a sloped power spectrum at low frequencies with an exponent *α* between 1 and 2. Variability of the fitted exponent for both short- and long-term evolution (Figure 1g) has been computed for all four patients over several sets of aDBS parameters.

### 4.3 Data preprocessing

We created several datasets according to the various predictions we performed, considering the two cases of a single set and multiple sets of aDBS parameters for the same patient. Each dataset contains LFP recordings of a single patient. They are organized automatically into three different files with beta power values (1-min resolution), amplitude of the spectrum in the 5-34 Hz range (10-min resolution), and stimulus current values delivered in both hemispheres (10-min resolution) respectively.

Starting from the device raw data, following the pipeline described in Supplementary Figure 3, we managed to create the Dataset needed within our LAURA framework where each entry is characterized by a tuple (*< input >, < label >*) where *< input >* consists of a tensor with the distributions of N consecutive days and *< label >* consists of a tensor with the distribution of the (N+K)th day. Importantly, daily beta power distributions have been min-max normalized to improve training efficacy. Specifically, for each patient we built three different datasets, according to the analysis performed:

1. Fix *K* = 1 (Figure 3a) such that for each entry we have: *< input >*= *tensor*([*distribution day*1, *distribution day*2, …, *distribution dayN*]) *< label >*= *tensor*([*distribution dayN* + 1]) with N varying from 1 up to 6. This first dataset contains entries derived from only one set of aDBS parameters.
2. Fix *N* = *N* ^*^ where *N* ^*^ is the optimal number of past days to perform a prediction (Figure 3b), resulted experimentally from the previous dataset (patients NWK1, NWK2: *N* ^*^ = 2, patients NWK3, NWK4: *N* ^*^ = 3) such that for each entry we have: *< input >*= *tensor*([*distribution day*1, *distribution day*2]) *< label >*= *tensor*([*distribution day*(2 + *K*)]) building several datasets according to the several values of K, with this parameter varying from 2 up to 6. All these datasets contain entries derived from only one set of aDBS parameters.
3. Same datasets as in 2. now derived from all the sets of aDBS parameters available for the same patient (Figure 3c), paying attention to the consecutiveness of the daily distributions for each entry.

Three copies of these three kinds of dataset have been built: one considering wholeday recordings and the other two differentiating between daytime and night recordings, to deal separately with the awake and asleep conditions. To allow the inter-patients generalization of the pre-processing pipeline used to build the dataset, few hyperparameters are needed:

- *driver channel* (either ‘Ch1’ or ‘Ch2’): hemisphere in which the power is monitored in order to choose the current to be delivered Ch1: left — Ch2: right
- (*Pwindow*_*min*_, *Pwindow*_*max*_): power range in which all the patient’s distributions are contained. According to our implementation, whatever interval between the two values would be divided into *n bins* = 206 bins.
- 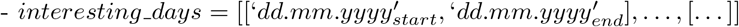: list of lists with [*start, end*] dates of the application of a specific set of aDBS parameters. The concept of ‘consecutiveness’ among days is applied only within the given intervals. The timeintervals of each patient are listed in Supplementary Table 2.

In all our experiments, we did not only consider the beta power distribution along all available recording hours, but also reduced the considered time intervals between 9:00am - 9:00pm in each day. This was of relevance as beta power can be impacted by sleep and therefore aDBS programming may be based by daily hours only.

### 4.4 Architecture

The proposed architecture aims to be implemented in the programming workflow to improve long-term DBS treatment for PD. Our aim is to predict a certain daily distribution of beta oscillatory activity based on recordings from previous days. This has very important clinical implications because it represents the first step toward IPGs that can automatically adjust stimulation parameters and set them based not on the immediate readout of the brain signal but on its evolution over time (see 3.Discussion).

We used a Transformer model (3.6 M parameters) to address our time-series forecasting problem, due to the multi-head attention (MHA) core, based on a general notion of sparsity of interactions within the sequence (Figure 2b). The input ‘*x*^′^ to the Transformer (see previous subsection) is a tensor of size [*N, n bins*] where *N* is the number of consecutive distributions used as input, *n bins* is considered as the embedding size. This means that each distribution in the sequence would represent a token, for which we can perform the one-step-ahead and the multi-step-ahead prediction. Given the input sequence, the Transformer processes it through three encoder layers and six decoder layers, followed by a FC layer with a sigmoid function as output non-linearity. The output of each layer for each position in the input sequence is a representation of that position after processing. When using Transformers for sequence-to-sequence tasks, the choice of *x*[− 1] as input for the FC layer stands for using the representation of the last position in the sequence as the final representation for the entire sequence. Since we are interested in predicting the distribution for the (*N* + *K*)th day, with *K* ∈ [1, 6], based on the information from the previous six days, using *x*[− 1] as input to the FC layer captures the model’s understanding of the entire sequence. The FC layer then projects this representation to the output space, with size *n bins*, to achieve the final prediction.

~~~
x <-input of size [batch\size, N, n\_bins]
x <-x + self.positional_encoding[:x.size(0), :]
x <-self.transformer(x, x)
x <-self.fc(x[:, -1, :])
return x
~~~

We need to set specific hyperparameters for our framework. We distinguish between dataset-related, architecture-related, and training-related hyperparameters, all listed in Supplementary Table 2 and Supplementary Table 3.

### 4.5 Training and Evaluation

We train the Transformer for non-autoregressive generation due to the sequential nature of the data in input. In this case the model learns to generate the output directly from the source sequence without considering previously generated tokens. That revealed as the best choice since autoregressive models generate each token conditioned on the previously generated tokens, meaning that errors or inaccuracies in predicting one token can propagate and affect subsequent predictions.

Several experiments have been performed on a single Quadro RTX 6000 GPU with 24 GB of Virtual RAM (VRAM). For each patient, three tasks have been investigated:

1. Task *N days* → 1 *dayafter* over a single set of aDBS parameters (Figure 3a), with N varying from 1 up to 6.
2. Task *N* ^*^ *days* → *K daysafter* over a single set of aDBS parameters (Figure 3b), with K varying from 2 up to 6, and where *N* ^*^ is the optimal number of past days to perform a prediction, resulted experimentally from the task 1.
3. Task *N* ^*^ *days* →*K daysafter* over multiple sets of aDBS parameters (Figure 3c), with K varying from 2 up to 6, and where *N* ^*^ is the optimal number of past days resulted from task 1.

Two different training strategies have been adopted accordingly to the case of single set (i, ii) or multiple sets (iii) of aDBS parameters. Each task has been performed for several values of the seed (ten seeds for the single set experiments, five seeds for the multiple sets experiments), randomly chosen, to initialize the NumPy, PyTorch, and PyTorchLightning libraries, in order to ensure robustness and reliability of the results despite the stochastic nature of deep learning techniques (i.e., random initialization of the network’s weights, data shuffling, dropout layers).

In the case where the data came from a single set, we have chosen that set for which we have more days of recording. Training was performed by splitting our dataset into two separate subsets, instead of the standard three subsets, due to the small number of entries extracted to deal with our tasks, containing 85% and 15% of the data for training and validation/test respectively. In order to prevent an increase in bias towards the validation/test set, no hyperparameter fine-tuning was conducted on this set. The number of epochs was set at 20000 independently of the experiments performed. As evidenced by the profiles of the training and validation losses, the model did not exhibit overfitting. Moreover, a crucial aspect of the training strategy is the choice of the criterion to measure the error between the observed distribution of the K th day and the predicted distribution for the same day. We used the *wMAPELoss*() criterion, which calculates the weighted mean absolute percentage error (wMAPE) between the true and predicted distributions, described as tensors. Such a weighted error allows different levels of importance or significance to be assigned to individual data points. By using this criterion, the means of the distributions are correctly estimated, the uncertainty around the mean is captured, and the zero values around the distributions are actually zero, all aspects that tend to be problematic when using other criteria such as *MSELoss*() (good mean estimation, very poor variance estimation) or the *MAPELoss*() (good mean and variance estimation, but with random selection of those values on either side of the distribution that are actually zero). In addition, *RMSProp*() was used as optimiser, with lr=1e-4 as the learning rate.

For the case where the data came from multiple sets of aDBS parameters, we trained our framework with a *quasi* − *LEAV E* − *ONE set*− *OUT*. This revised version of the LEAVE-ONE-OUT technique consists of two steps: 1) training the model on several sets of aDBS parameters, leaving one set out; then, since the model goes in overfitting, 2) we perform a fine tuning on the left out set of parameters, training the framework on a train set with only 50% of the data and then testing it on the remaining data. The set left out for each patient is the second one for which we have more days of recording, in order to achieve more reliable evaluation results. This way, overfitting is solved, resulting in comparable performance to the previous tasks. We used *wMAPELoss*() as criterion, with RMSProp() as optimizer and lr=1e-5 as learning rate.

For the sake of clarity, we outline our pipeline for distinguishing between training and validation sets:

- Single Set of aDBS parameters:
  1. Partition recordings from a single patient based on different aDBS parameter settings.
  2. Form training/validation tuples: For one specific set of aDBS parameters, segment daily recordings into tuples (*x, y*), e.g., ([*day*_*n′*_, *day*_*n′*_+1], *day*_*n′*_+*k*).
  3. Split the dataset: Assign 85% of the data to training and 15% to validation, then shuffle the training set.
- Multiple Sets of aDBS parameters:
  1. Partition recordings from a single patient based on different aDBS parameter settings.
  2. Form training tuples for each parameter setting as described above.
  3. Two-phase validation approach:
  – Phase 1: Exclude one set of aDBS parameters, using its tuples as the validation set while using all other sets for training. The training set is shuffled.
  – Phase 2: Within the previous validation set, allocate 50% of the data for finetuning and 50% for final validation. The fine-tuning subset is shuffled.

In this way, we avoid LAURA learning dependencies between the daily distributions presented during validation/test during training, thus mitigating overfitting (Supplementary Figure 8).

Dealing with a distributional regression task, several metrics have been computed on the prediction, such as RMSE, MAPE, R2, and wMAPE. We assess the performance of our framework by computing the average wMAPE, over the bins of the predicted distributions, over the test set. Searching for bounded metrics for regression tasks, we also computed the average over the test set of the percentage inaccuracies in identifying the 10th and 90th percentiles of the predicted distributions with reference to the observed ones. This metrics have been chosen strictly correlated to the thresholds *β*_*min*_, *β*_*max*_ settled during the aDBS device recalibration.

### 4.6 Linear and Zero-order predictor

To demonstrate LAURA’s effectiveness in predicting beta power variations, we compared LAURA’s performance with that of a linear regressor and that of a zero-order regressor, as defined below.

Given a sequence of time series data *source sequence* with length *num days*, we fit a simple linear regression model to each bin of the time series such that:

~~~
next value = a * (current value) + b
~~~

For *num days* = 1, we set a zero order regressor with *a* = 1 and *b* = 0; For *num days* = 2, the model directly computes the coefficients from the last two days; For *num days >* 2, the model uses linear interpolation across more days to estimate coefficients.

Let’s denote:

- *x*_*t*_ = [*x*_*t*1_, *x*_*t*2_, …, *x*_*tn*_]^*T*^ as the data vector for day t, where *n* = *num*_*b*_*ins*;
- *m* = *num days*;
- *a*_*i*_ and *b*_*i*_ as the coefficients for the linear model for bin *i*.

The linear model for predicting the data for bin *i* at day *t* + 1 based on the data at day *t* is:

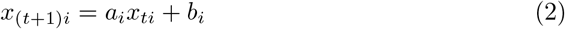

To fit this linear model for each bin i, we need the following matrices:

- the Design Matrix *X* such that *X*[*i*, :] = [*x*_1*i*_, *x*_2*i*_, …, *x*_(*m*−1)*i*_] is a row vector containing all data for bin *i* from day 1 to day (*m* − 1).
- the Target Matrix *Y* such that *Y* [*i*, :] = [*x*_2*i*_, *x*_3*i*_, …, *x*_*mi*_] is a row vector containing
- the next day’s data for bin *i*, thus from day 2 to day *m*.

Finding the coefficients *a*_*i*_ and *b*_*i*_ that best fit the linear relationship between *X*[*i*, :] and *Y* [*i*, :] means to solve the following LS (least square) optimization problem for bin i:

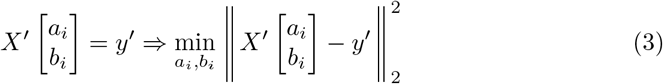

where *X*^′^ = [*X* [*i*, :]^*T*^ 1], *y*^′^ = *Y* [*i*, :]^*T*^. The solution to the LS optimization problem is deterministic:

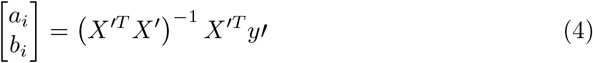

When *num days* = 2, we only have a linear model based directly on the two days for each bin:

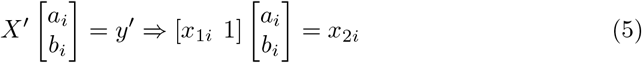

If *x*_1*i*_ ≠ 0, then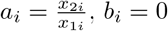.

If *x*_1*i*_ = 0, then *a*_*i*_ = 0, *b*_*i*_ = *x*_2*i*_.

### 4.7 Random Forest Algorithm (RFA) predictor

As further confirmation of the inherent nonlinearity of the subthalamic beta activity, we compared piecewise linear and zero order regressors with a Random Forest Algorithm (RFA) to predict one day ahead. Since it does not make any a priori assumption on the dynamics, the RFA is able to capture non-linearities, yet it does not inherently have an understanding of the time sequence (that’s why only the one-day-ahead prediction is performed).

According to the notation introduced in Section 4.6, for each bin *i* of the the input distribution, we fit a Random Forest model such that:

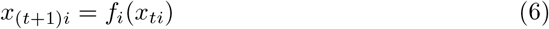

where *f*_*i*_ is trained on historical data.

The Random Forest regressor is an ensemble of *R* decision trees. Each tree makes a prediction *h*_*r*_(*x*_*ti*_), and the final prediction is the average of all tree predictions. Our RFA implementation leverages the `RandomForestRegressor` from the scikit-learn library in Python.

## Data Availability

The datasets presented in this article are not readily available because LFP recorded with the AlphaDBS device cannot be deposited in a public repository as they can be traceable to the identity of the subject. Requests to access the datasets should be directed to IUI.

## Data and Code availability

Python code for the LAURA framework will be made publicly available upon acceptance of this manuscript.

## Acknowledgments

The author(s) declare that financial support was received for the research, authorship, and/or publication of this article. The study was funded by the European Union - Next-Generation EU - NRRP M6C2 - Investment 2.1 Enhancement and strengthening of biomedical research in the NHS, and by the Fondazione Pezzoli per la Malattia di Parkinson. CP and IUI were supported by the Deutsche Forschungsgemeinschaft (DFG, German Research Foundation) Project-ID 424778381 - TRR 295. IUI was supported by a grant from New York University School of Medicine and The Marlene and Paolo Fresco Institute for Parkinson’s and Movement Disorders, which was made possible. With support from Marlene and Paolo Fresco. AM was supported by the Italian Ministry of Research, under the complementary actions to the NRRP “Fit4MedRob - Fit for Medical Robotics” Grant (# PNC0000007). AM was supported by the project “IMAD23ALM MAD: The etiopathological basis of gait derangement in Parkinson’s disease: decoding locomotor network dynamics”. SF and AM acknowledge the support of the BRIEF “Biorobotics Research and Innovation Engineering Facilities” project (Project identification code IR0000036) funded under the National Recovery and Resilience Plan (NRRP), Mission 4 Component 2 Investment 3.1 of Italian Ministry of University and Research funded by the European Union – Next-Generation EU.

We are grateful to many colleagues for their help in this line of research. In particular, we would like to thank: Salvatore Bonvegna, Elena Contaldi, and Manuela Pilleri of the Parkinson Institute of Milan, ASST G.Pini-CTO; Nicoló Pozzi and Ibrahem Hanafi of the University Hospital of Würzburg; Lorenzo Piangatello and Michael Lassi of the Biorobotics Institute of the Sant’Anna School of Advanced Studies.

## Author contributions

SF: Conceptualization, Formal Analysis, Investigation, Methodology, Software, Writing–original draft. LC: Conceptualization, Data curation, Investigation, Methodology, Writing–review and editing. CB: Data curation, Writing–review and editing. CP: Conceptualization, Data curation, Investigation, Methodology, Writing–review and editing, Funding acquisition, Resources, Supervision. IUI: Conceptualization, Formal Analysis, Investigation, Methodology, Writing–original draft, Funding acquisition, Resources, Supervision. AM: Conceptualization, Formal Analysis, Investigation, Methodology, Writing–original draft, Funding acquisition, Resources, Supervision.

## References

[1] Engel, A. K. & Fries, P. Beta-band oscillations—signalling the status quo? Current opinion in neurobiology 20, 156–165 (2010).

[2] Kühn, A. A. et al. Pathological synchronisation in the subthalamic nucleus of patients with parkinson’s disease relates to both bradykinesia and rigidity. Experimental neurology 215, 380–387 (2009).

[3] Neumann, W.-J. & Kühn, A. A. Subthalamic beta power-unified parkinson’s disease rating scale iii correlations require akinetic symptoms. Mov Disord 32, 175–176 (2017).

[4] Priori, A., Foffani, G., Rossi, L. & Marceglia, S. Adaptive deep brain stimulation (adbs) controlled by local field potential oscillations. Experimental neurology 245, 77–86 (2013).

[5] David, F. J., Munoz, M. J. & Corcos, D. M. The effect of stn dbs on modulating brain oscillations: consequences for motor and cognitive behavior. Experimental brain research 238, 1659–1676 (2020).

[6] Johnson, K. A. et al. Proceedings of the 11th annual deep brain stimulation think tank: pushing the forefront of neuromodulation with functional network mapping, biomarkers for adaptive dbs, bioethical dilemmas, ai-guided neuromodulation, and translational advancements. Frontiers in Human Neuroscience 18, 1320806 (2024).

[7] Cometa, A. et al. Clinical neuroscience and neurotechnology: An amazing symbiosis. Iscience 25 (2022).

[8] Marceglia, S. et al. Deep brain stimulation: is it time to change gears by closing the loop? Journal of Neural Engineering 18, 061001 (2021).

[9] Little, S. et al. Bilateral adaptive deep brain stimulation is effective in parkinson’s disease. Journal of Neurology, Neurosurgery & Psychiatry 87, 717–721 (2016).

[10] Stanslaski, S. et al. Sensing data and methodology from the adaptive dbs algorithm for personalized therapy in parkinson’s disease (adapt-pd) clinical trial. npj Parkinson’s Disease 10, 174 (2024).

[11] Isaias, I. U. et al. Case report: Improvement of gait with adaptive deep brain stimulation in a patient with parkinson’s disease. Frontiers in Bioengineering and Biotechnology 12, 1428189 (2024).

[12] Caffi, L. et al. Adaptive vs. conventional deep brain stimulation: One-year subthalamic recordings and clinical monitoring in a patient with parkinson’s disease. Bioengineering 11, 990 (2024).

[13] Arlotti, M. et al. Eight-hours adaptive deep brain stimulation in patients with parkinson disease. Neurology 90, e971–e976 (2018).

[14] Reich, M. M. et al. Progressive gait ataxia following deep brain stimulation for essential tremor: adverse effect or lack of efficacy? Brain 139, 2948–2956 (2016).

[15] St. George, R., Nutt, J., Burchiel, K. & Horak, F. A meta-regression of the long-term effects of deep brain stimulation on balance and gait in pd. Neurology 75, 1292–1299 (2010).

[16] Lachenmayer, M. L. et al. Subthalamic and pallidal deep brain stimulation for parkinson’s disease—meta-analysis of outcomes. NPJ Parkinson’s disease 7, 77 (2021).

[17] Canessa, A., Palmisano, C., Isaias, I. U. & Mazzoni, A. Gait-related frequency modulation of beta oscillatory activity in the subthalamic nucleus of parkinsonian patients. Brain Stimulation 13, 1743–1752 (2020).

[18] Avantaggiato, F. et al. Intelligibility of speech in parkinson’s disease relies on anatomically segregated subthalamic beta oscillations. Neurobiology of Disease 185, 106239 (2023).

[19] Vissani, M. et al. Impaired reach-to-grasp kinematics in parkinsonian patients relates to dopamine-dependent, subthalamic beta bursts. npj Parkinson’s Disease 7, 53 (2021).

[20] LeCun, Y., Bengio, Y. & Hinton, G. Deep learning. nature 521, 436–444 (2015).

[21] Thenaisie, Y. et al. Towards adaptive deep brain stimulation: clinical and technical notes on a novel commercial device for chronic brain sensing. Journal of Neural Engineering 18, 042002 (2021).

[22] Golshan, H. M., Hebb, A. O. & Mahoor, M. H. Lfp-net: A deep learning framework to recognize human behavioral activities using brain stn-lfp signals. Journal of neuroscience methods 335, 108621 (2020).

[23] Yang, Y. et al. Modelling and prediction of the dynamic responses of large-scale brain networks during direct electrical stimulation. Nature biomedical engineering 5, 324–345 (2021).

[24] Cook, M. J. et al. Prediction of seizure likelihood with a long-term, implanted seizure advisory system in patients with drug-resistant epilepsy: a first-in-man study. The Lancet Neurology 12, 563–571 (2013).

[25] Karoly, P. J. et al. Forecasting cycles of seizure likelihood. Epilepsia 61, 776–786 (2020).

[26] Aiello, G., Valle, G. & Raspopovic, S. Recalibration of neuromodulation parameters in neural implants with adaptive bayesian optimization. Journal of Neural Engineering 20, 026037 (2023).

[27] Wernisch, L. et al. Online bayesian optimization of vagus nerve stimulation. Journal of Neural Engineering 21, 026019 (2024).

[28] Michmizos, K. P., Sakas, D. & Nikita, K. S. Prediction of the timing and the rhythm of the parkinsonian subthalamic nucleus neural spikes using the local field potentials. IEEE Transactions on Information Technology in Biomedicine 16, 190–197 (2011).

[29] Khawaldeh, S. et al. Subthalamic nucleus activity dynamics and limb movement prediction in parkinson’s disease. Brain 143, 582–596 (2020).

[30] Abbes, M. et al. Subthalamic stimulation and neuropsychiatric symptoms in parkinson’s disease: results from a long-term follow-up cohort study. Journal of Neurology, Neurosurgery & Psychiatry 89, 836–843 (2018).

[31] Haddad, A. R., Samuel, M., Hulse, N.Lin, H.-Y. & Ashkan, K. Long-term efficacy of constant current deep brain stimulation in essential tremor. Neuromodulation: Technology at the Neural Interface 20, 437–443 (2017).

[32] Ruge, D. et al. Shaping reversibility? long-term deep brain stimulation in dystonia: the relationship between effects on electrophysiology and clinical symptoms. Brain 134, 2106–2115 (2011).

[33] Schade, S., Mollenhauer, B. & Trenkwalder, C. Levodopa equivalent dose conversion factors: an updated proposal including opicapone and safinamide. Movement disorders clinical practice 7, 343 (2020).

[34] Vaswani, A. Attention is all you need. Advances in Neural Information Processing Systems (2017).

[35] Heaton, J. Ian goodfellow, yoshua bengio, and aaron courville: Deep learning: The mit press, 2016, 800 pp, isbn: 0262035618. Genetic programming and evolvable machines 19, 305–307 (2018).

[36] Ge, Y. et al. Openagi: When llm meets domain experts. Advances in Neural Information Processing Systems 36 (2024).

[37] Najafabadi, M. M. et al. Deep learning applications and challenges in big data analytics. Journal of big data 2, 1–21 (2015).

[38] Whang, S. E., Roh, Y., Song, H. & Lee, J.-G. Data collection and quality challenges in deep learning: A data-centric ai perspective. The VLDB Journal 32, 791–813 (2023).

[39] Jin, P., Lu, L., Tang, Y. & Karniadakis, G. E. Quantifying the generalization error in deep learning in terms of data distribution and neural network smoothness. Neural Networks 130, 85–99 (2020).

[40] Rossi, E., Rosa, M., Rossi, L., Priori, A. & Marceglia, S. Webbiobank: A new platform for integrating clinical forms and shared neurosignal analyses to support multi-centre studies in parkinson’s disease. Journal of Biomedical Informatics 52, 92–104 (2014).

[41] Caré, M., Chiappalone, M. & Cota, V. R. Personalized strategies of neurostimulation: from static biomarkers to dynamic closed-loop assessment of neural function. Frontiers in Neuroscience 18, 1363128 (2024).

[42] Tang, S., Modi, A., Sjoding, M. & Wiens, J. Clinician-in-the-loop decision making: Reinforcement learning with near-optimal set-valued policies. International Conference on Machine Learning 9387–9396 (2020).

[43] Bakken, S. Ai in health: keeping the human in the loop. Journal of the American Medical Informatics Association 30, 1225–1226 (2023).

[44] Zhou, H. et al. A survey of large language models in medicine: Progress, application, and challenge. arXiv preprint 2311.05112 (2023).

[45] Marceglia, S. et al. Double-blind cross-over pilot trial protocol to evaluate the safety and preliminary efficacy of long-term adaptive deep brain stimulation in patients with parkinson’s disease. BMJ open 12, e049955 (2022).

