## Supplementary Materials for "Transformer-based long-term predictor of subthalamic beta activity in Parkinson’s disease"

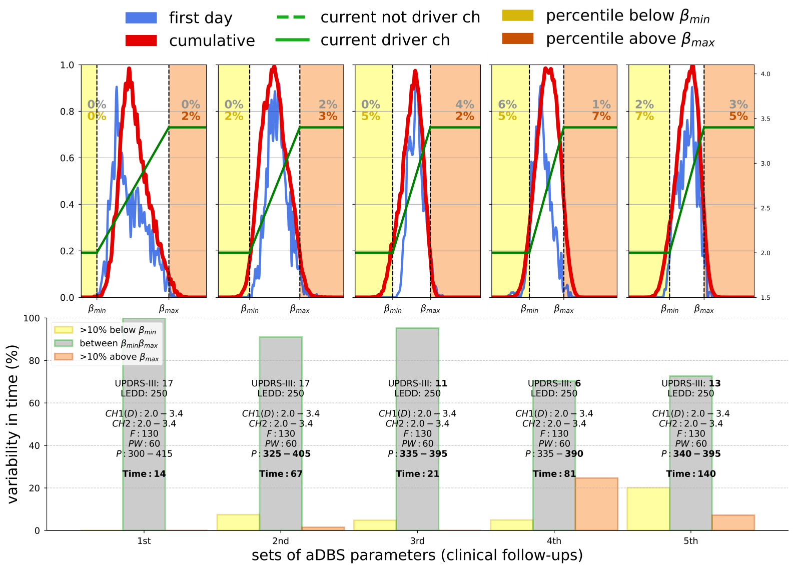


(a) Patient NWK1

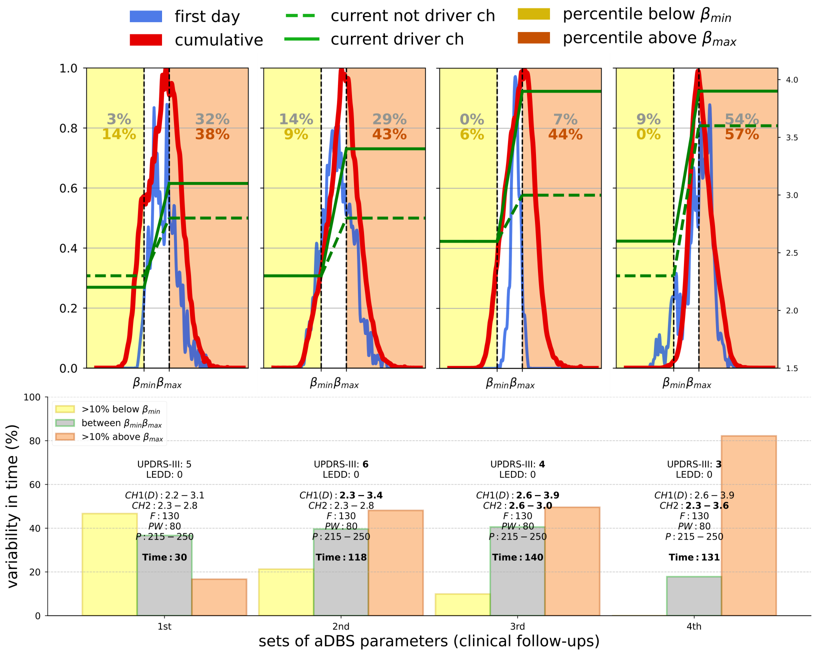


(b) Patient NWK2

**Supplementary Fig. 1**: **Explorative Data Analysis (EDA) for all patients.** The figure is similar to Figure 1 in the main text for each of the four patients (a: NWK1, b: NWK2, c: NWK3, and d: NWK4) with the only difference being that the daily distribution of patient-specific beta power is reported as cumulative of all days *(continues...)*

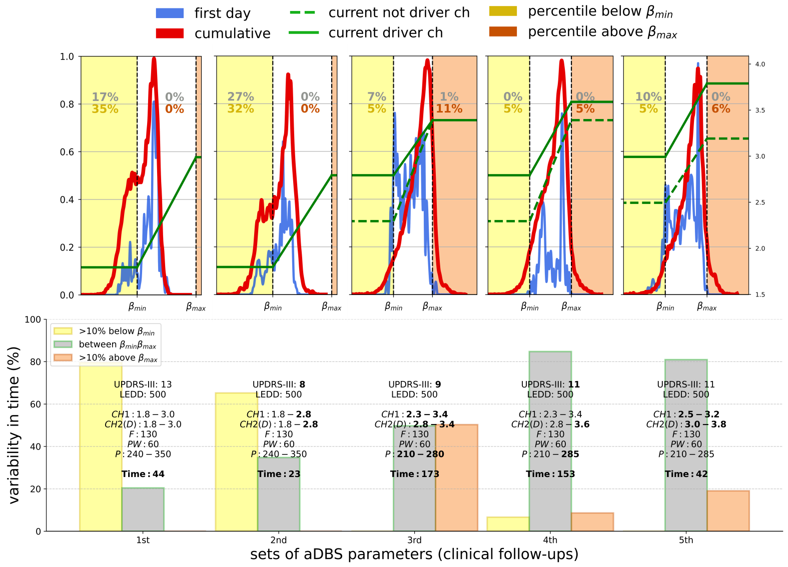


(c) Patient NWK3

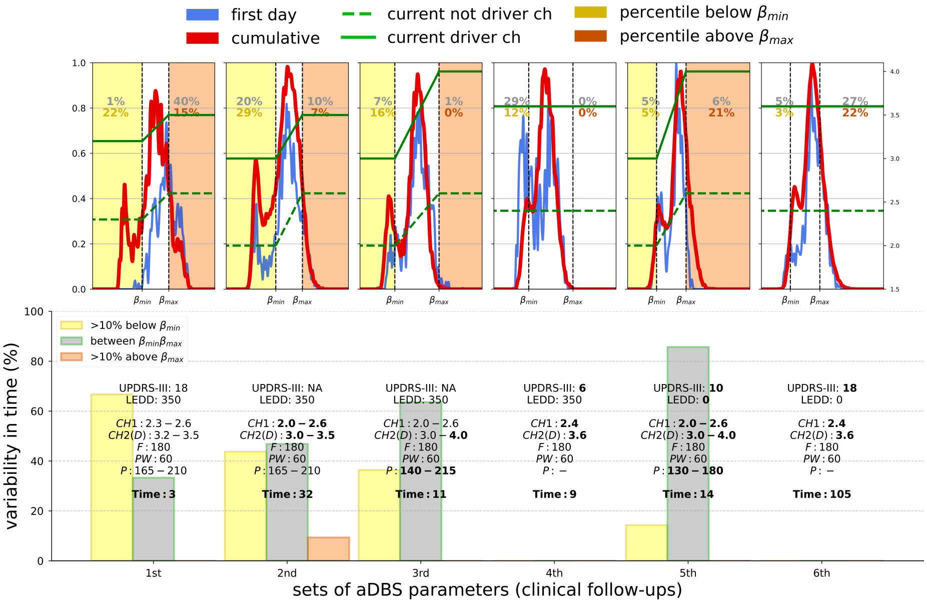


(d) Patient NWK4

**Supplementary Fig. 1**: **Explorative Data Analysis (EDA) for all patients.** *(continues...)* prior to a visit (red line, upper panel) and as the first day after aDBS reprogramming (blu line, upper panel). For patient NWK4, at the 4th and 6th visit DBS was set in conventional mode (cDBS). For these panels, the $\beta_{min}$ and $\beta_{max}$ are set to the previous values for visual comparisons but were not used for DBS programming.

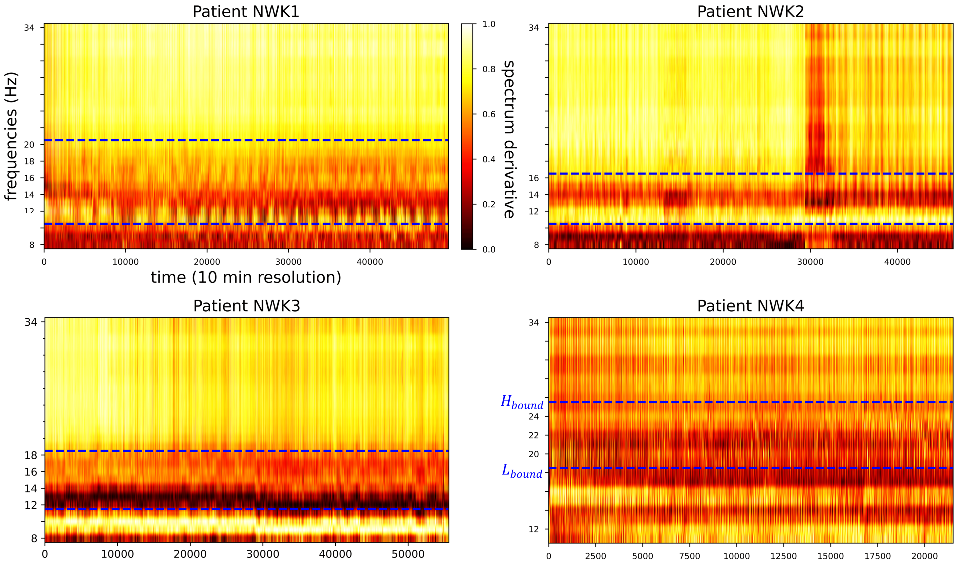


**Supplementary Fig. 2**: **Stability of the beta peak over time.** The power spectrum (PS) derivative, extracted from the driver channel, is visualized for each patient as a coloured map. The frequency peak is clearly visible and remained stable over time within the patient-specific beta frequency range (dashed blue lines identify the Lbound and Hbound, see Table 1). The power measured between Lbound and Hbound was used as input signal for the linear proportional algorithm for aDBS. For the sake of the colour coding, the original frequency range [5, 34] Hz has been cut appropriately for each patient.

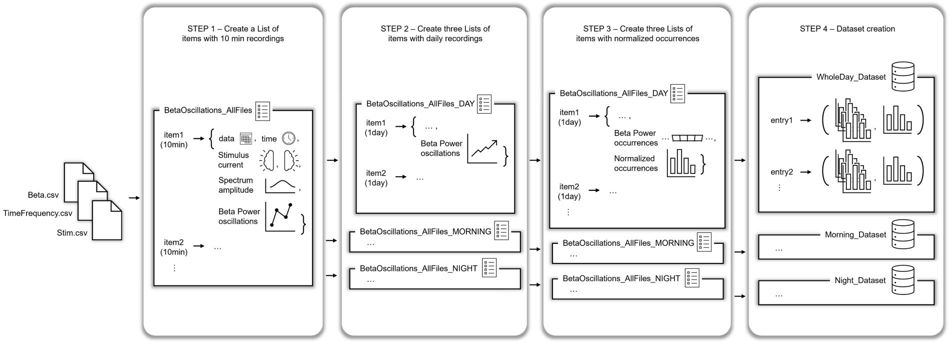


**Supplementary Fig. 3**: **Dataset generation.** The dataset needed for the training of LAURA in a supervised-learning approach is generated directly from the data received by the AlphaDBS device. STEP1: a list of dictionaries is created such that each dictionary represents a recording of 10 min (consisting of 1 sample of power spectrum, 1 sample of the stimulus current, 10 samples of the beta power oscillations in the time domain). STEP2: three different lists of dictionaries are generated according to the time stamps of each item on the list of step1. Each dictionary represents a recording of 1 whole day or 12h day-time or 12h night, according to the list generated. STEP3: from each item of the lists generated in step2, the daily beta power distributions are extracted and min-max normalized according to improve training efficacy. STEP4: several datasets are generated according to the day-ahead to predict and the length of the history sequence.

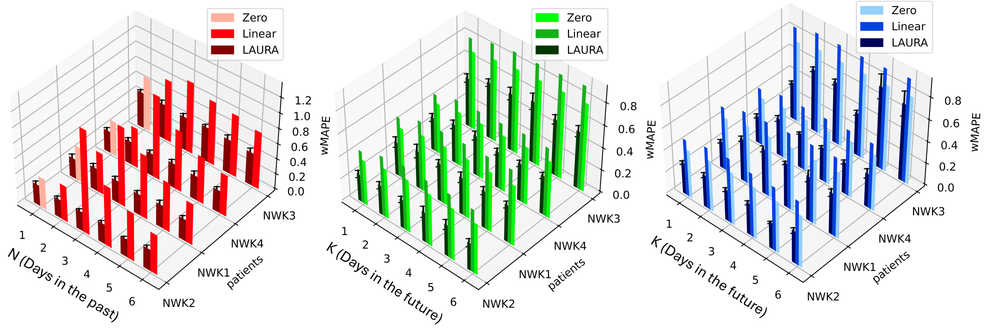


**Supplementary Fig. 4**: **Patient-dependent performance of LAURA for all subjects.** Left (red): one-step-ahead prediction over a single aDBS set. Centre (green): multi-step-ahead prediction over a single aDBS set. The wMAPE is monitored on predictions from 1 up to 6 days ahead, based on the patient's history sequence of optimal length N*. Specifically, N*=2 days for patients NWK1 and NWK2, while N*=3 for patients NWK3 and NWK4. Right (blue): multi-step-ahead prediction over multiple aDBS settings. The wMAPE is monitored on predictions from 1 up to 6 days ahead, based on the patient's history sequence of optimal length N*.

**Supplementary Table 1**: Demographic and clinical characteristics

| | **NWK1** | **NWK2** | **NWK3** | **NWK4** |
|---|---|---|---|---|
| Gender (M or F) | M | M | M | M |
| Age (years) | 50-60 | 40-50 | 50-60 | 30-40 |
| Disease duration at last recordings (years) | 15-20 | 10-15 | 15-20 | 10-15 |
| Stimulating contacts (Left/Right) | 2-C+ / 9-C+ | 1-C+ / 8-C+ | 2-C+ / 9-C+ | 2-C+ / 9-C+ |
| Recording contacts | 1-3 | 0-2 | 8-10 | 10-11 |
| Beta frequency range ([Lbound-Hbound] Hz) | 11-20 | 11-16 | 12-18 | 19-25 |

* Quadripolar 3389 leads (Medtronic) have the following contacts enumeration rule: 0 or 8 are the ventralmost contacts, 3 or 11 are the dorsalmost contacts, respectively for the left and right STN.
Age and duration of disease are reported in decades to preserve anonymity.

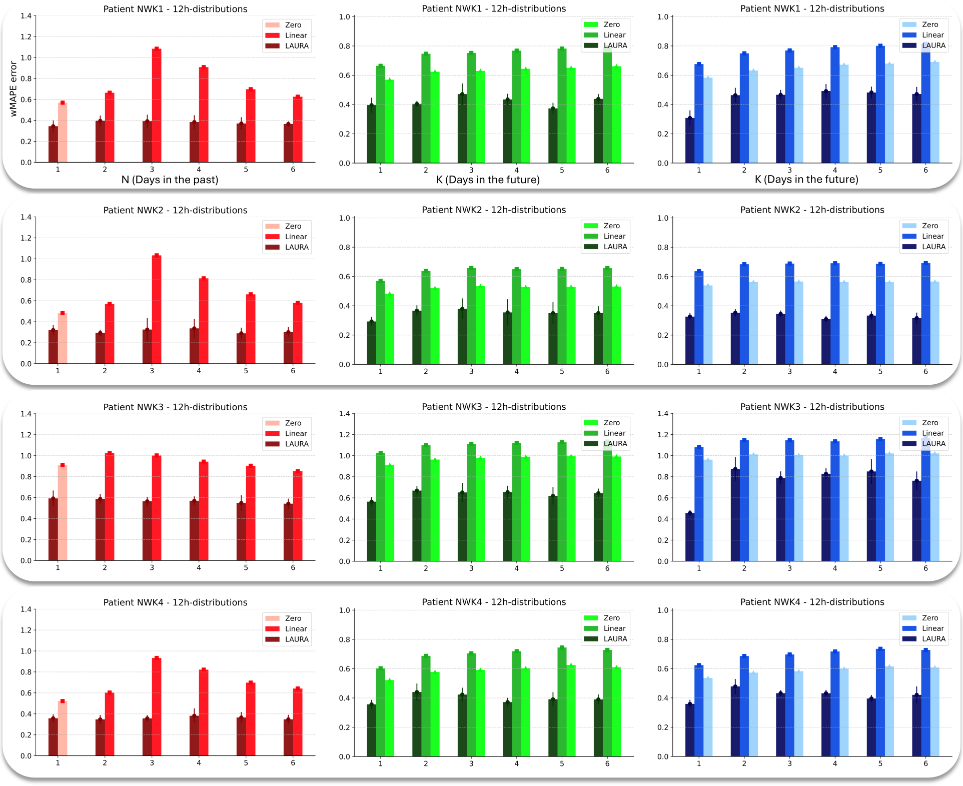


**Supplementary Fig. 5**: **Patient-dependent performance of LAURA for all patients on 12h distributions.** The performance of LAURA based on 12-hour (9:00am-9:00pm) distribution is comparable to that of 24-hour distribution. Evaluation results are shown for all patients. Specifically, N*=2 days for patients NWK1 and NWK2, while N*=3 for patients NWK3 and NWK4. Left (red): one-step-ahead prediction over a single aDBS set. Centre (green): multi-step-ahead prediction over a single aDBS set. Right (blue): multi-step-ahead prediction over multiple aDBS settings.

**Supplementary Table 2**: Data inspection. Four patients: NWK1, NWK2, NWK3, NWK4.

| Dataset Hyperparameters | NWK1 | NWK2 | NWK3 | NWK4 |
|---|---|---|---|---|
| Driver channel (CH1: left; CH2: right) | CH1 | CH1 | CH2 | CH2 |
| $Pwindow_{min}$ (in a.u.) | 276.0 | 149.0 | 90.0 | 81.0 |
| $Pwindow_{max}$ (in a.u.) | 482.0 | 355.0 | 341.0 | 287.0 |
| Distribution length (in bins) | 206 | 206 | 206 | 206 |
| Time evaluated (in days)*** | **NWK1:** 18, 44, 14, 67, 21, 81**, 140*<br>**NWK2:** 30, 53**, 17, 22, 9, 6, 30, 24, 52, 116*, 13<br>**NWK3:** 44, 23, 173*, 153**, 42<br>**NWK4:** 3, 32**, 11, 9, 14, 105* | | | |

* aDBS setting analysed in the single set experiments
** aDBS setting left out in the multiple sets experiments
*** The total number of days used for actual training/testing may differ from those shown in Figures 1 and Suppl. Figure 1. This is due to sporadic missing days in long-term recordings within a fixed aDBS parameters period, which we separated.

**Supplementary Table 3**: Hyperparameters from LAURA (3.6 M Params)

| Parameter | Value |
|---|---|
| **Architecture Hyperparameters** | |
| Batch size | 32 |
| Input size | 206 |
| Num. of head attention | 2 |
| Hidden size | 256 |
| Num. layers Encoder | 3 |
| Num. layers Decoder | 6 |
| **Training Hyperparameters** | |
| **- Single set of aDBS parameters** | |
| Seeds | 1101, 1203, 1504, 2106, 2206, 2408, 2504, 2903, 2907, 3101 |
| Criterion | wMAPELoss() |
| Optimizer | RMSProp |
| Learning rate | 1e-4 |
| Num. of epochs | 20k |
| **- Multiple sets of aDBS parameters** | |
| Seeds | 1101, 1203, 2106, 2408, 3101 |
| Criterion | wMAPELoss() |
| Optimizer | RMSProp |
| Learning rate STEP1 | 1e-5 |
| Learning rate STEP2 | 1e-5 |
| Num. of epochs STEP1 | 20k |
| Num. of epochs STEP2 | 10k |

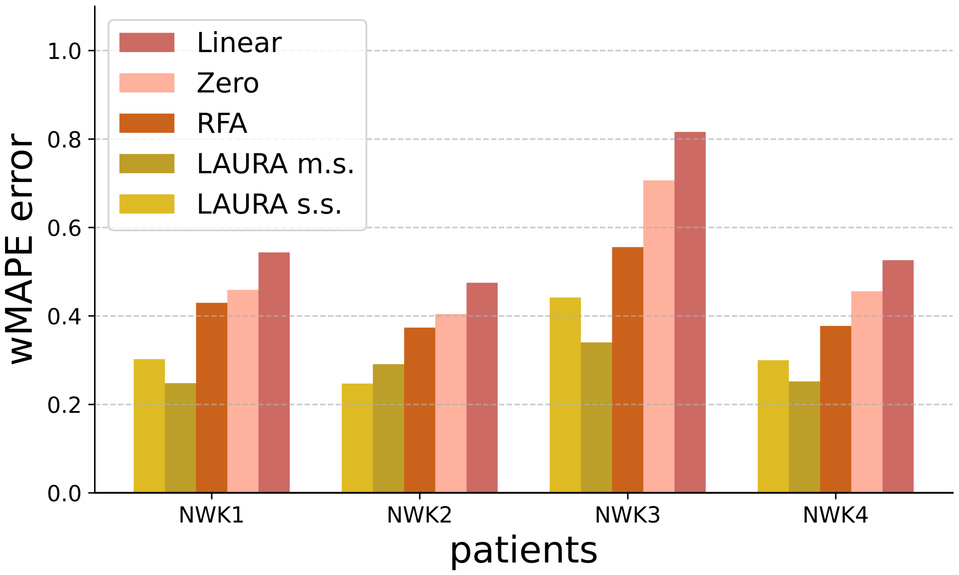


**Supplementary Fig. 6**: **Comparison of zero-order, piecewise linear, and non-linear regressors.** We provide further confirmation of the inherent non-linearity of subthalamic beta power activity by comparing for each patient the prediction error of (from right to left for each patient) the piecewise linear regressor, the zero-order regressor, the RFA regressor, LAURA in the multi-setting scenario, and LAURA in the single-setting scenario. The one-day-ahead predictions were compared. The RFA regressor outperforms the linear regressors. LAURA outperforms all other regressors.

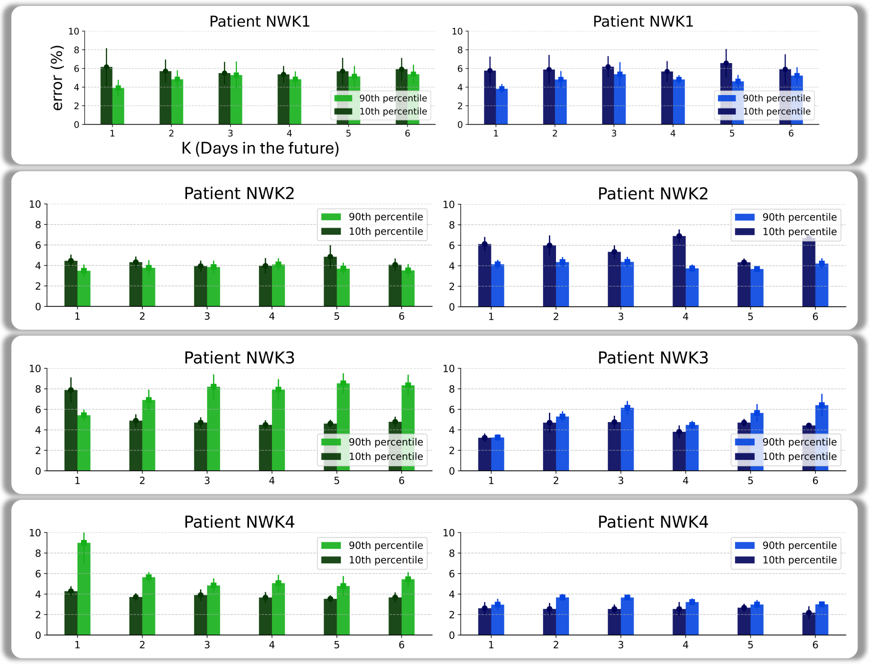


**Supplementary Fig. 7**: **Functional evaluation of predicted distributions with aDBS for all subjects.** We show the evaluation of the inaccuracies of LAURA in identifying the 10th (dark) and 90th (light) percentile of each daily distribution, for single (left) and multiple (right) aDBS programming sets. The results promote LAURA as a patient-specific system.

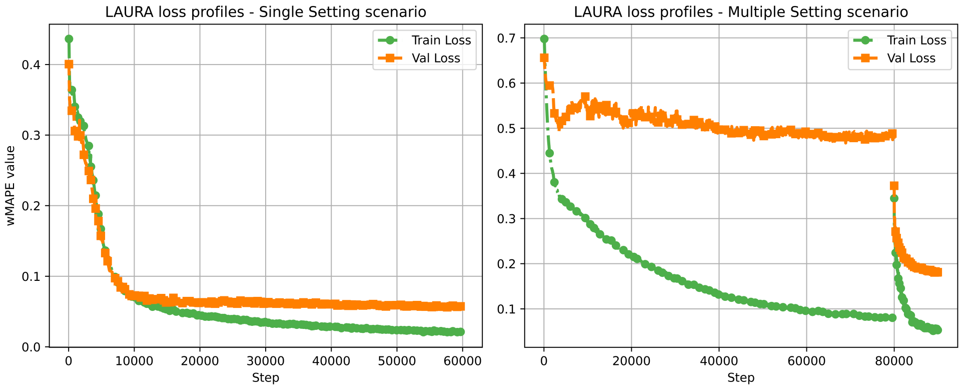


**Supplementary Fig. 8**: **Plots of (representative) Loss profiles during the LAURA training process.** We monitored the training (Train Loss, in green) and the validation (Val Loss, in orange) losses associated with the LAURA's Transformer model. (Left) Single Setting scenario, patient NWK4 - one-day-ahead prediction. (Right) Multiple Setting scenario, patient NWK4 - one-day-ahead prediction.
